# On predicting the novel COVID-19 human infections by using Infectious Disease modelling method in the Indian State of Tamil Nadu during 2020

**DOI:** 10.1101/2020.04.05.20054593

**Authors:** Arsath Abbasali Ayubali, Sara Roshini Satheesh

**Affiliations:** Department of Mechanical Engineering, College of Engineering Guindy, Anna University Chennai – 600 025, Tamil Nadu, India; Independent Researcher, Social Anthropologist, Chennai – 600 008, Tamil Nadu, India

**Keywords:** Novel Corona Virus 19 (COVID-19), Cluster analysis, Epidemiology, Infectious Disease modelling (IDM), Travellers, Risk, Basic reproduction number, SEIR Model and modified SEIR Model

## Abstract

Since the introduction of the novel Corona Virus (The COVID-19) to the Chinese city Wuhan in the Hubei province during the late December 2019, the effectiveness of the deadly disease, its human infection, spreading severity and the mortality rate of the infection has been an issue of debate. The outbreak of the virus along the time has become a massive threat to the global public health security and has been declared as a pandemic. Accounting the radical number of increases in the infected cases and the death due to COVID-19 infections around the globe, there is a need to predict the infections among the people by making proper optimization and using various Infectious Disease modelling (IDM) methods, in order to challenge the outcome. In comparison with previous diseases like SARS and Ebola viruses, the new corona virus (COVID-19) infections are infectious during the incubation period. In addition to that, naturally produced droplets from humans (e.g. droplets produced by breathing, talking, sneezing, coughing) and Person-to-person contact transmission are reported to be the foremost ways of transmission of novel corona virus. By considering the above two factors, a modified SEIR (Susceptibility-Exposure-Infection-Recovery) method have been used for predicting the spread of the infections in the state of Tamil Nadu which is located in the southern part of India. Further, we have utilized the current surveillance data from Health and Family Welfare Department – Government of Tamil Nadu to accurately predict the spreading trend of the infection on a state level.

## 1. Introduction

Since the Novel Corona Virus 19 (COVID-19) was first detected in Hubei province of Chinese country in Wuhan City in late December 2019, it has been spreading rapidly across the globe (European Centre for Disease Prevention and Control, 2020; Tang, Zhou., Li, Xianbin., Li, Houqiang., 2020). Furthermore, by the end of March 2020, COVID-19 had been detected in most of the countries globally leading a pandemic situation (Tang, Zhou., Li, Xianbin., Li, Houqiang., 2020). Nevertheless, the COVID-19 was declared as a global pandemic by The Director General of the World Health Organization (WHO) on 11 March 2020.

Subsequently, as of 15 March 2020, 1 69 517 cases of COVID-19 were reported worldwide by more than 150 countries. Since late February, the majority of cases reported are from outside China, with an increasing number of these reported mainly from the USA, Italy. Spain and from most part of EU/EEA countries and the UK (Tang, Zhou., Li, Xianbin., Li, Houqiang., 2020; WHO, 2020). As the number of infections among the globe increases, several thousands of cases are reported by EU/EEA countries to be dead due to the severe illness caused by the COVID-19 infections. Further, on March 25^th^, the world report (WHO, 2020) on corona virus infections reported that, Italy represents 15 % of the infected cases (74,386) and 35 % of the fatalities (7,503) of the total infected and dead cases reported around the globe. The current hop of the increase in cases around the EU/EEA and the UK reflects the trends which had been realised in China during the January and early February 2020. In the present situation where COVID-19 infections are hastily spreading worldwide and as initially the number of cases in Europe were rising with an increasing pace in several affected areas, but now a days due to the infected travellers moving around the globe for various activities are spreading the infections to the persons who come in near contact with the infected individuals. Due to such mass transmission of the infections, there is a need for instantaneous targeted action towards the spread of COVID-19. The speed with which COVID-19 can cause nationally devastating epidemics once transmission within the community is established (European Centre for Disease Prevention and Control, 2020) which specifies that may be in a few weeks or even days, it is most likely that situations to those seen in China, USA and Italy may be seen in other states of India.

Significantly, at this current situation there are no proven medicines for curing the COVID-19 or vaccines to prevent the transmission of the infections but instead there are only very few evidences on the effectiveness of potential therapeutic treatments to improve the immune system of an individual as a precaution. The clinical demonstrations of COVID-19 range from no symptoms (asymptomatic) to respiratory problems and lastly to severe pneumonia; severe infections can induce multi-organ failure and lead to death (WHO, 2020). The majority of reported cases (80%) are with milder respiratory infections and pneumonias. It is noticed that severe illness and deaths are more common among the ageing population with other chronic pre-existing conditions. These are the risk groups which are accounting for the majority of severe disease and fatalities to date (Rhonda, Sue., Roberts., Ivo, M Foppa., 2006).

Previously, several pandemics such as SARS (severe acute respiratory syndrome), Ebola, HIV/AIDS, pandemic influenza has been reported to be originated in animals which are caused by viruses, and are determined to develop by behavioural, environmental conditions, or socioeconomic fluctuations (Stephen, S Morse., Jonna, A. K. Mazet., Mark, Woolhouse., Colin, R. Parrish., Dennis, Carroll., William, B. Karesh., Carlos, Zambrana-Torrelio., 2012). The 2009 influenza A (H1N1) pandemic was one of the most closely tracked and studied epidemics in history (Manoj, G., Catherine, B., Justin, J., Amra, U., Lucinda, E. J., Matthew, B., 2015). The source of the new coronavirus (COVID-19) has yet not been identified and proven scientifically though several researches had been conducted for past one decade and more by various research scholars and scientists. Several reports (European Centre for Disease Prevention and Control, 2020; WHO, 2020) claims that the source of the infection of the COVID-19 comes from wild animals. Indeed, majority of the research works indicate that, it is most analogous to a corona virus family isolated from bats and the transitional host may be pangolin, yet it is still uncertain that which kind of wild animal may be the source.

As stated above, the human-to-human transmission of the new coronavirus is mainly caused by respiratory droplets, physical touch of the infected areas and close contact with the infected persons. From the clinical reports from various regions, it is evident that the incubation period of the COVID-19 virus might be from 3-14 days and also it is infectious even during the incubation period, which is the serious cause of the increased number of infections emerging around the globe.

In spite of the nation’s considerable effects on public health and increased understanding of the process by which the infections emerge, no pandemic has been forecasted before infecting human beings. Previously, reports have been reviewed about what is known about the pathogens that emerge, the hosts from which the specific infections originate in, and the factors that drive their transmission (Stephen, S Morse., Jonna, A. K. Mazet., Mark, Woolhouse., Colin, R. Parrish., Dennis, Carroll., William, B. Karesh., Carlos, Zambrana-Torrelio., 2012). On the one hand, various epidemiologist have discussed the challenges to control their spread and new efforts to predict pandemics and identify prevention strategies (Ogden, Nicholas H., Fazil, Aamir., Safronetz, David., Drebot, Michael A., Wallace, Justine., Rees, Erin E., Decock, Kristina., Ng, Victoria., 2017). Apparently, having such innovative technologies and facilities such as new mathematical modelling, diagnostic, communications systems, and informatics technologies, one can identify and forecast the spread along with its risk of infections and make approaches that are needed to control and safeguard the public health (Ogden, Nicholas H., Fazil, Aamir., Safronetz, David., Drebot, Michael A., Wallace, Justine., Rees, Erin E., Decock, Kristina., Ng, Victoria., 2017).

On the other hand, traditional epidemiological forecasting methods, such as laboratory-based surveillance and outbreak investigations were swiftly used to advise policy decisions makers during the pandemic situations (“Outbreak of 2009 pandemic influenza A (H1N1) at a school—Hawaii, May 2009,”; Han, K., Zhu, X., He, F., 2009; Lessler, J., Reich, N. G., Cummings, D. A., Nair, H. P., Jordan, H. T., Thompson, N., 2009; Luliano, A. D., Reed, C., Guh, A., 2009). Concurrently, extensive contributions were made in the study of infectious disease modeling (IDM) (Cauchemez, S., Bhattarai, A., Marchbanks, T. L., 2011; Cauchemez, S., Donnelly, C. A., Reed, C., 2009; Mostaco-Guidolin, L. C., Bowman, C. S., Greer, A. L., Fisman, D. N., Moghadas, S. M., 2012; Presanis., Pebody, R. G., Paterson, B. J., 2011; Towers, S., Feng, Z., 2009). The purpose of this article could be a guide to the way in which one of the Infectious Disease Modelling (IDM) method contributes to policy deliberations and decision-making in grounding for, or when experiencing a pandemic situation.

As the COVID-19 is not an Indian born virus infection to human being and it has origin in China, the infection to the Tamil Nadu population must only via transmission, specifically through the travelers who are already infected of COVID-19 within the incubation period of 3-14 days. The possibility of infection in Tamil Nadu (TN) region also depends on the total numbers of travellers and the proportions that use air travel to reach any part of Tamil Nadu region since the initial discovery of the disease. Therefore, in order to manage the risks of residents of Tamil Nadu, a modified SIER method (one of the IDM methodology) is chosen to predict the number of infected cases of COVID-19 by applying the current available data from the Ministry of Health, Tamil Nadu, India.

## 2. Methodology

There are extensive and eminent histories of mathematical models in epidemiology. Since the ancient times, theoretical epidemiology has been witnessing numerous developments in various directions. Some of these studies can be found in (Bailey, 1975), (Anderson, R. M., May, R. M., 1991) and (Hethcote, 2000). An incredible number of models have been framed, analyzed and applied to an assortment of infectious diseases qualitatively and quantitatively. In addition, mathematical models have been the most significant tools in analyzing the spread and to determine the control measures of the infectious diseases. Additionally, mathematical models currently play a key role in the areas of policy making, including health-economic aspects, socio-economic aspects, emergency planning and risk valuation, spreading control evaluation, and optimizing various aspects.

Almost all of the mathematical models in predicting the epidemiology is based on the Compartmental Models which is a technique used to simplify the mathematical modelling of infectious disease. In this model the population is divided into compartments, with the assumption that every individual in the same compartment has the same characteristics (Sarah A, 2012).

As a significant element, SEIR model is presented in this paper, where there is an exposed duration between being infected and becoming infective (Incubation period). Various researches have been conducted on SEIR models which are presented in detail in (Sun, C., Hsieh, Y., 2010; Yi, N., Zhang, Q., Mao, K., Yang, D., Li, Q., 2009; Zhang, J., Li., Ma, Z., 2006; Zhou, X., Cui, J., 2011). Treating such deadly disease such as SARS, measles, flue and tuberculosis this model played a significant role in controlling or decreasing the spread of the infections in the specified study region. More recent work on the effect of treatment on the dynamic behavior can be found in (Tang, Zhou., Li, Xianbin., Li, Houqiang., 2020; Wang, J., Liu, S., Zheng, B., Takeuchi, Y., 2012; Wendi, 2006). Contrastingly, in traditional epidemic models, the treatment rate is presumed to be proportionate to the number of infective, which is almost not possible in reality. Therefore, in this paper assuming that the virus-infected person is not infectious during the incubation period and that the infected person did not take isolation measures during the illness, will not be suitable for the current situation as the COVID-19 is infectious even during its incubation period. Hence, this article first describes,

1. A modified SEIR model.
2. Predict and analyze the varying trend of the epidemic situation.
3. Estimate the parameters involved in the infection dynamics model.

Furthermore, from outputs of the above considerations, using Matlab to simulate the established dynamic equations from the model which will be based on recent public data from the Ministry of Health - Tamil Nadu and analyze the results. Also further providing recommendations for local prevention and control of the COVID-19 infectious diseases.

### 2.1 The Modified SEIR Model

The novel Corona virus infection when compared with SARS, the symptoms of virus infection are latent in patients which are obvious from the reports of the confirmed patients. Therefore, this quality brings great complications for the prevention and control of the epidemic. Form the various reports and analysis, one infers the classical SEIR model assumes that the infected person’s incubation period is not infectious; this assumption will not be valid for this case of COVID-19 infection characteristics. Therefore, a modified or improved SEIR model is considered to analyze and predict the trend of the epidemic.

The modified SEIR model is similar to the traditional SEIR model by amending the following assumptions:

1. The normal birth rate and mortality during the study period are not taken into account;
2. The influence of other peripheral factors on the model parameters is overlooked during the study period;
3. The transmission method is considered as from infected human-to-human.

For predicting the virus infections due to human-to-human transition in the Tamil Nadu state, the total population of the state has to be divided into four categories:

> *Susceptible populations* (S, not yet infected but at risk of being infected),
>
> *Exposed or latent people* (E, including people with mild or asymptomatic but infectious populations),
>
> *Infected* (I, confirmed to be infectious) and *Recovered* (R, including those who have been cured and died from COVID-19 infection).

The first case of Novel Corona virus infection was found on March 7, 2020 in the State Tamil Nadu. It is reported from quarantine and clinical observations (WHO, 2020), that the average incubation period of New Corona virus infection was 7 days, so considering that, one believes that the first patient who is with infection of COVID-19 may already exist on February 29, 2020. It is said that, the infected patient has a travel history to Oman. In order to obtain a precise prediction, we have classified those who have been infected with the COVID-19 but are not diagnosed as latent human; when the latent humans are diagnosed as COVID-19 infected case, those will be treated in a special quarantine hospital. This type of conclusion will lead to the situation that, infected people will not be having the possibility of spreading and transmitting the virus to the others. For these reasons, we aim to predict the rate of infections in the south Indian region state of Tamil Nadu by using a modified SEIR model described above. The parameters of the SEIR model, which are dependent on time function ‘*t*’ and are modelled as below,

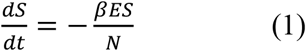

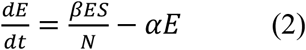

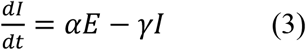

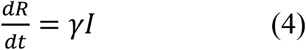

Where

*N*,corresponds to the total population which represents *N* = *S* + *E* + *I* + *R*

*β*,indicates (referred to as the effective contact rate) the probability of infection between the susceptible and the latent, which is expressed as *β* = *k* * *b*, where *k* in the latent population daily exposed and *b* is the probability of the population being infected

*α*,represents the conversion rate of the latent human to the infected human, which is the obtained as the reciprocal of the incubation duration

*γ*,denotes the recovery rate, which is computed by the reciprocal of number of days the infected human is in treatment.

Additionally, the death parameter is included in the model to predict the number of deaths for the considered period of prediction. According to the reports from WHO (World Health Organisation) (WHO, 2020), the death rate has been estimated as 0.034 based on the upcoming clinical reports and previous incidents. Therefore, the death parameter has been formulated as below,

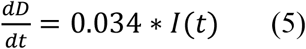

Further, in this analysis, there is a consideration that the person infected with the novel Corona Virus 2019 has an incubation period during which the infected person could transmit the virus to a healthy person during this incubation period. These kinds of transmission during the incubation period have not been reported in case of Ebola or SARS viruses. Therefore, the rate of change in the states of the SIER parameter will depend on the past state and not only on the current state. Specifically, during the analysis of the modified SIER model, the present number of infections (*I*(*t*)) which depends on the time constraints is considered as two fragments. One was transferred by the human carrying the incubating COVID-19 virus and the other is the number of confirmed humans who are infected with the virus at the time interval ‘*t*’. A recent report (Qun, Li., M, Med., 2020) has assessed the incubation period and the average duration of the incubation period of the COVID-19 is reported to be 5.2 days (95 % of the Clinical Investigations are reported to be between 4.1 to 7 days) (Qun, Li., M, Med., 2020). Therefore, in this analysis, for an efficient analysis, the incubation period has been chosen to be 7 days. Hence, from the above consideration, at a sample time ‘*t*’ an infected person who is confirmed to be infected with COVID-19 would have been exposed to the virus from a latent person before 7 days of the confirmation report. Consequently, from the assumptions made above, the modified SEIR model can be constructed using delay difference in the parameters,

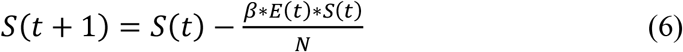

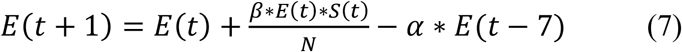

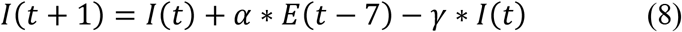

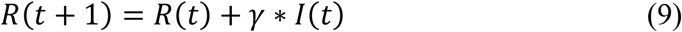

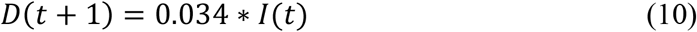

### 2.2 Analysis of the model parameters

In this analysis, the situation which has prevailed in the Chinese city of Wuhan and the outcome reports is considered as the base model. Therefore, while mapping the analysis to Tamil Nadu, the parameters have to be assigned with the data which will be in accordance with the current statistical data of Tamil Nadu region. According to the census report 2012 of Government, the population of Tamil Nadu was officially recorded as 67.86 million. Therefore, certain projected studies reveal that the population of Tamil Nadu in 2019 was 77, 177, 540 (The Director General, 2012). Therefore, in this analysis it is assumed that N = 77, 177, 540. When a susceptible human (S) is getting contact with that latent human (E) in a unit time ‘*t*’, the susceptible human can be infected with the virus having the most probability of getting infected. As a result, that latent human becoming a possible virus carrier (E).

According to clinical investigations and official reports (WHO, 2020), from 2 % to 5 % of adjacent contacts of the infected or latent human are diagnosed as infected. Therefore, the probability of contact between a susceptible human and a latent human is from 2 % to 5 %. In this analysis we use the maximum, which is 5 % as the probability of infection ‘*b*’. The average daily number of the close contacts of the latent person is *k* = 5. Therefore, the probability of infection ‘*β*’ is 0.25 (*β* = *k* * *b*).

In the current situation each of the exposed human (E) is transformed into an infected human (I) at a conversion rate ‘*α*’, and it is assumed from the reports (WHO, 2020) that, the incubation period refers to the duration from the infection acquired by the human to the outcome of clinical symptoms of the virus. From the reports of WHO (WHO, 2020), COVID-19 is initially assessed to have an incubation duration of 2 to 12.5 days and a median duration of 5 to 6 days. Although, most of the recent reports (Tang, Zhou., Li, Xianbin., Li, Houqiang., 2020) indicates that, the incubation period for COVID-19 may be up to or little more than 14 days from the initial infection of the virus. Hence, WHO recommends a quarantine period of 14 days with constant follow-up of the symptoms. Therefore, in this analysis, it is assumed that the incubation period is a maximum 7 days. Therefore, the conversion rate of the latent human to the infected human *α* = 0.14 which is *α* = 1/(*incubation period*). The probability of each infected human (I) becoming a recovered human (R) is duration of the treatment process. The treatment process can vary from each individual, but in this analysis the earliest time of recovery recorded is taken into account. It is evident from reports (Tang, Zhou., Li, Xianbin., Li, Houqiang., 2020) that, the treatment plan is premeditated to allow the infected person to continue 14 days under quarantine and attain an auto healing effect through the infected human’s autoimmune mechanism. Therefore, the treatment cycle selected in this paper is 9.1 days based on the various clinical reports (WHO, 2020) and the Wuhan model, so the recovery rate is 0.11 (*γ* = 1/9.1 = 0.11).

From the above analysed conclusions, the parameters for the modified SIER model have been assumed. The parameters *S*(*t*),*E*(*t*),*I*(*t*), and *R*(*t*) are all greater than zero. As per the ministry’s report the first patient in Tamil Nadu region was found on March 7, 2020. Therefore, the first case of latency would have appeared on February 29, 2020. Now, from the above considerations February 29, 2020 is the 0th day, so the modified SIER parameters at ‘*t* = 0’ will be *S*(0) = 77,177,539, *E* (0) = 1, *I* (0) = 0, and *R* (0) = 0.

The above formulations and parameters have been subjected to analysis by using Matlab software. The outcome of the results and its inferences are discussed in the sections below.

## 3. Results and Discussion

By using Matlab software two models have been simulated. The one which is based on the real time predictions, where there are no full influential governmental policies enacted to the public and the other is, where the government has proposed various actions for the welfare of the people and to prevent the spread of the COVID-19 infections. Therefore, both the models have been simulated using Matlab and the current available data.

While simulating the modified SIER model before enacting policies, we can get the figure 1. Day 0 in Figure 1 corresponds to the time when the first human infection case (latency duration) was found, that is, February 29, 2020. It can be seen from Figure 1 that the epidemic situation starts to concentrate in about 90 days. It can be seen that the number of infected people will reach its peak in about 120 days.

**Figure 1:**
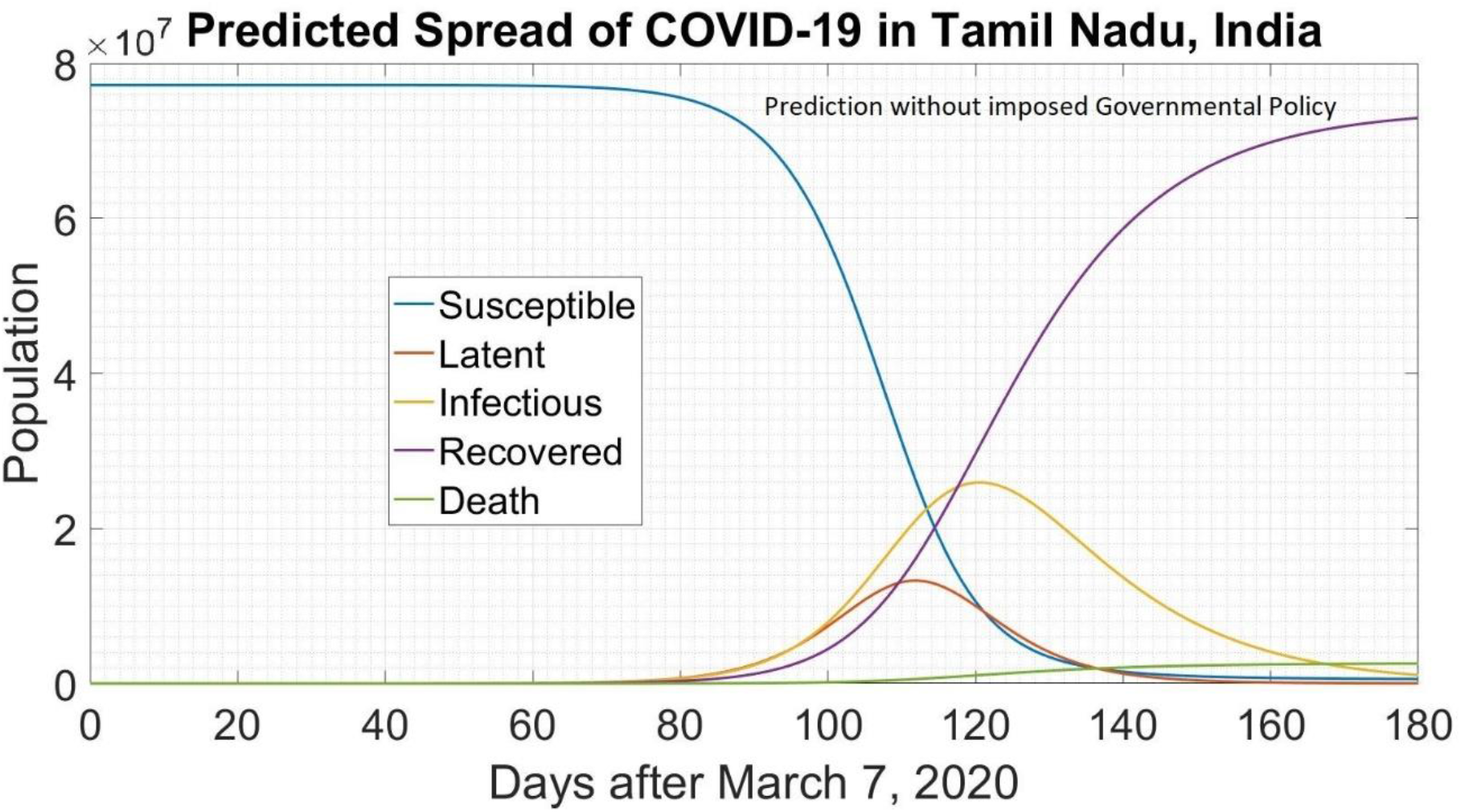
Prediction plot of COVID-19 in Tamil Nadu, India without Governmental policies.

From the above analysis the Table.1 shows the number of Infected, Latent and the dead humans during the prediction period from March 7, 2020.

**Table 1:**
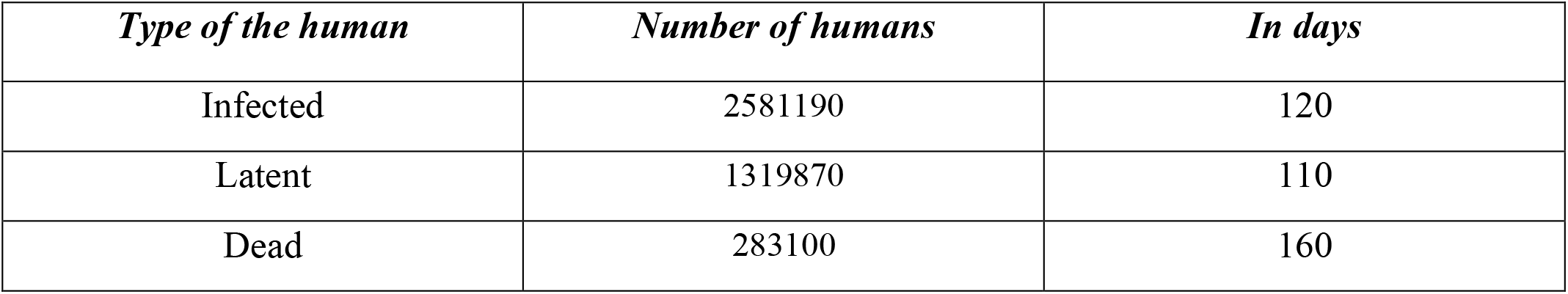
Prediction of the number of Infected, Latent and Dead in Tamil Nadu, India without Governmental policies

Secondly, while considering the second case of prediction in the case of enacted governmental policies like quarantine precautions, hand sanitizing, social distancing and avoiding the crowd in public places, the Figure. 2 shows the predicted results. From the predicted results, it is obvious that, the number of infected people may reach 13, 24 and 470 only in 220 days and will eventually come to an end after 250 days from the period of initial consideration.

**Figure 2:**
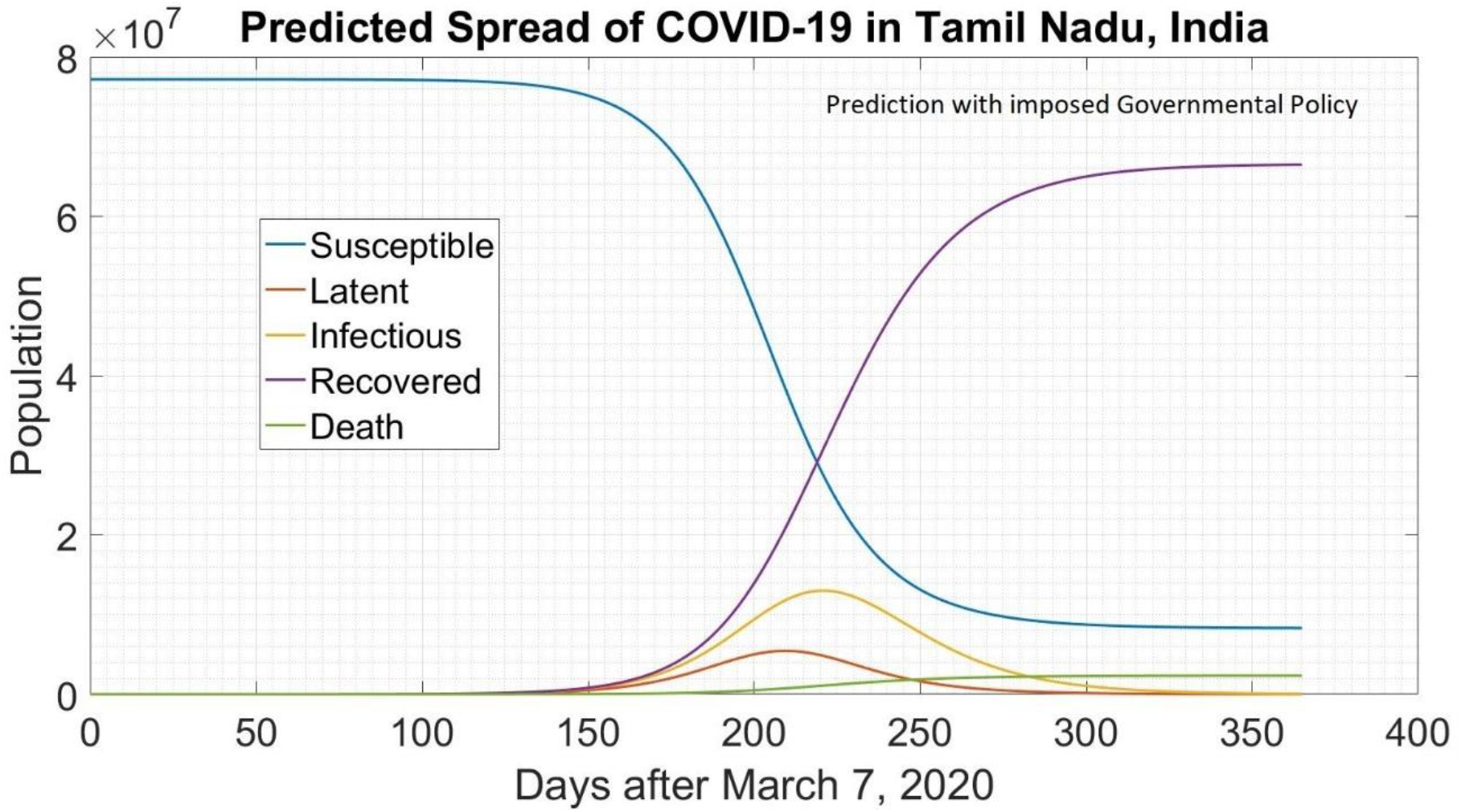
Prediction plot of COVID-19 in Tamil Nadu, India with imposed Governmental policies.

In actual fact, seeing Wuhan’s model, its Municipal Government had not only implemented the city closure measures on January 23, but also regulated traffic facilities in Wuhan and closed down most public places. The similar effect has been considered in the simulated model by reducing the contact ration to half. Therefore, the change in the number of infected persons are reduced which can be obtained in Figures. 2. It is obvious from Figure. 2 that, when the government takes strong precautionary measures to control the average daily number of contacts per infected person, the number of infected persons reaches a peak after about 220 days, and the number of infected persons is reduced to 13, 24 and 470. It is also evident from Table. 2 that, when the government adopts effective quarantine measures, the number of infected people can be drastically reduced.

**Table 2:** Prediction of the number of Infected, Latent and Dead in Tamil Nadu, India with imposed Governmental policies

| *Type of the human* | *Number of humans* | *In days* |
| --- | --- | --- |
| Infected | 1324470 | 220 |
| Latent | 529660 | 210 |
| Dead | 166181 | 250 |

## 4. Conclusions

On the one hand, since February 21, 2020, all the countries across the globe have been adopting strict isolation and precautionary measures, and on the other hand, Wuhan has closed all public transportation facilities, air travel, parks and almost a complete lock down since January 20, 2020. From the previous news articles and the managements reports, China’s response to the epidemic can be seen in three stages: first stage is to control the primary source of infection and discard the transmission chains around all the areas in Hubei Province; second stage is to vigorously treat infected patients and reduce mortality; and the third stage is to control the epidemic condition, thoroughly contain the disease epidemic, and drastically reduce the contact ratio between human population. In this article, the path of the curve is simulated by a time-lapse modified SEIR model. The model prediction shows that without any governmental policies or control measures, the peak of infected human will reach about 25, 81, 190 when the average daily number of contacts per infected person is *k* = 5.

Therefore, it is recognised that the Tamil Nadu Government and the Ministry of Health takes strict control and precautionary measures from March 22, 2020. Hence, the number of people infected will be greatly reduced which is expected to be by the simulated model as 13, 24 and 470 of the total population of Tamil Nadu. Therefore, local government authorities and public health departments should emphasise their efforts on controlling the source of infection and delink the maximum possible transmission chains, such as increasing the sample number of testing which leads to the early detection of the infected person, screening of the latent humans which finds its way to early isolation, and early preparedness for treating the infected humans expected to come, to highly cut down the gathering of large groups of people, and to regularly and completely ban the use of public toilets and public utility areas. Importantly the policies are to me made in such a way that, it should strictly impose on the “Three Cs” - Closed and Crowded places and Conversations in close proximity to the public thereby creating a awareness to avoid the community transmission of the COVID-19.

## Data Availability

The data sets generated during and/or analysed during the current study are available from the corresponding author on reasonable request.

## Acknowledgement

The authors wish to thank various people in the Government Office, Ministry of Health, Government of Tamil Nadu for their help in collecting the data. Special thanks should be given to Prof. K. A. Padmanabhan and Prof. Emeritus. Dr. Josephus Platenkamp for their professional guidance and valuable support in building the author’s research and presentation abilities.

